# Water Treatment and E. Coli in Drinking Water: Household Responses to (Invisible) Water Quality Risks

**DOI:** 10.1101/2025.08.14.25333747

**Authors:** Akito Kamei, Bhowmik Sujey Soori

## Abstract

**Background:** In 2024, an estimated four billion people lacked access to safely managed drinking water, with the greatest risks in low- and middle-income countries, particularly for vulnerable groups such as young children and pregnant women.

**Methods:** This paper analyzes data from 59,633 households in 25 countries across Sub-Saharan Africa, Latin America, and Asia to examine E. coli contamination in source water and household responses through water treatment. It investigates whether households are more likely to treat water when source water is contaminated and how treatment relates to E. coli levels in stored drinking water.

**Findings:** The study finds that 78 percent of households do not treat their water, while 20 percent rely on sources with high E. coli contamination (*>*100 MPN/100ml). The presence of E. coli is associated with a modest increase in water treatment—typically three to five percentage points, depending on the contamination level—driven by greater use of chlorine products or straining/settling methods. While water treatment is linked to lower contamination levels in stored drinking water, a large share of households that treat their water still face moderate to high E. coli contamination risk at the point of consumption.

**Interpretation:** These findings highlight that the key challenge is not only whether households treat their water, but whether they do so properly and consistently. Observational evidence from real-world conditions points to the need for more WASH strategies that go beyond promoting access to treatment, ensuring its correct application to deliver safe drinking water.

**Funding:** No funding was received for this research.

**Research in context:** *Evidence Before This Study:* Previous research on water quality and health in low-income countries has primarily focused on access to water infrastructure or small-scale trials of water treatment interventions. Only recently have nationally representative surveys begun testing for microbial contamination at the point of use within households. Analyses of data from the UNICEF Multiple Indicator Cluster Surveys (MICS) have shown that E. coli contamination is widespread, even in improved water sources, highlighting the need for effective household-level treatment (Husein, Nounkeu, Armah, & Dharod, 2023; Joseph, Haque, Moqueet, & Rong Hoo, 2019; Kandel, Kunwar, Lamichhane, & Karki, 2016). Using evidence from 27 countries, Bain, Johnston, Khan, Hancioglu, and Slaymaker (2021) demonstrate that contamination frequently persists at the point of consumption. This study is among the first to examine household water treatment behavior in direct relation to measured E. coli contamination, using large-scale, nationally representative data.

*Added value of this study:* This study offers a large-scale multi-country analysis of drinking water safety and household responses, using data from 59,633 households in 25 countries in Sub-Saharan Africa, Latin America, and Asia. In contrast to research that relied on proxy indicators of contamination, this study uses direct microbiological testing of source and drinking water for E. coli contamination at a large scale. By pooling multiple nationally representative DHS surveys, the study achieves broad external validity. The paper documents the share of households exposed to contamination at the source and the proportion who treat their water among those at risk, as well as how water treatment practices relate to lower levels of E. coli in stored drinking water. This framework offers new, policy-relevant evidence grounded in real-world conditions. It advances our understanding of the proportion of people treating water, among those exposed to high water contamination risk, and how effectively these practices result in safe drinking water.

*Implications of all the available evidence:* Despite well-documented health risks from waterborne pathogens (Nataro & Kaper, 1998), treat-ment adoption remains low even among highly exposed households. Furthermore, the weak relationship between existing treatment methods and E. coli-free drinking water highlights the need for more scalable and effective WASH interventions. The collective evidence underscores that ensuring safe drinking water in LMICs requires not only improving access to water sources but also promoting and sustaining effective water treatment at the household level. The findings from this study, in line with prior knowledge, highlight that many households remain exposed to fecal contamination because they do not consistently treat their water, do not have the means to measure the contamination level, or because treatment is not always effective. This implies that water, sanitation, and hygiene (WASH) policies must go beyond simply encouraging water treatment behavior: they should ensure that households have the means, knowledge, and motivation to treat water properly and consistently. By emphasizing correct and routine use of household water treatment, alongside improvements in source water quality, public health programs can more effectively protect families from waterborne illnesses and advance progress toward truly safe drinking water for all.

## 1 Introduction

In 2024, an estimated four billion people globally lacked access to safely managed drinking water, with vulnerable populations like young children, pregnant women, and those with weak immune systems facing heightened risks from contamination (Hope, 2024). This challenge is particularly acute in low- and middle-income countries (LMICs), where water contamination is widespread. In Africa, the estimated number of people without safe drinking water rose from 703 million in 2015 to 766 million in 2021 (UN Water, 2021). Waterborne pathogens, including fecal contamination from *Escherichia coli (E. coli)*, pose significant health risks, especially in regions where contamination persists (Black & Talbot, 2005).

This paper uses nationally representative Multiple Indicator Cluster Surveys (MICS) data from 59,633 households across 25 countries in Sub-Saharan Africa, Latin America, and Asia to document E. coli contamination in source water and examine how households respond to its invisible risk through water treatment, as well as how water treatment practices relate to lower level of E. coli in stored drinking water. We begin by documenting the share of households that treat their water. Next, we report the share of households exposed to contamination at the water source and the share that treat their water among those exposed to some risk of E. coli. We then empirically estimate whether the likelihood of water treatment increases based on the level of source water contamination. Finally, after presenting E. coli contamination in stored drinking water, we report how contamination levels change from source to point of consumption, comparing households that treat their water to those that do not.

On average, 34 percent of households across the 25 countries rely on water sources with moderate contamination (1–100 MPN/100ml), and 20 percent rely on sources with high contamination levels (*>*100 MPN/100ml).^1^ Despite this widespread exposure, only 22 percent of households report treating their water. Empirical analysis, controlling for factors such as country fixed effects and water source type, finds that moderate contamination is associated with a three-percentage-point increase in the likelihood of water treatment, while high contamination corresponds to a five-percentage-point increase. This pattern holds across all regions, with varying magnitudes of the coefficients. The increase is particularly evident for treatment methods such as chlorination (including Aquatabs and PUR) and straining or settling, but not for boiling.

The contamination level of stored drinking water at the point of consumption is higher than that of the water at the source, with 28.4 percent of drinking water samples showing high levels of contamination (*>*100 MPN/100ml). When the household does not treat the water, the share of households exposed to high contamination rises from 20 percent at the source to 34 percent in the stored drinking water.

Water treatment is associated with lower E. coli levels in drinking water, but complete elimination remains rare. Even among households that treat their water, 15 percent of those who boil, 33 percent who use chlorine, and 28 percent who strain or settle water still face high levels of contamination. Only 55 percent of households that boil their water, 21 percent of those using chlorine, and 17 percent of those who strain or settle their water have drinking water free of E. coli.

While these are not causal estimates of water treatment effectiveness—for instance, households using chlorine or straining/settling often start with more contaminated source water than those who boil or do not treat—the analysis offers valuable insights into how well different treatment practices succeed, or fall short, in achieving safe drinking water, based on observational data from nationally representative surveys. This suggests that proper application and post-treatment storage practices are just as important as the decision to treat water itself.

This paper makes three contributions to the existing WASH literature. First, it documents how households adjust water treatment behaviors in response to the invisible contamination risks. Prior work has shown that perceptions, more than objective risk, often drive household decisions (Manski, Charles F., 2004), and access to improved water sources may even cause households to reduce water treatment efforts (Bennett, 2008; Jalan & Ravallion, 2003; Jessoe, 2013). Since E. coli contamination is invisible, households may incorrectly believe their water is safe (G. Schuitema, T. Hooks, & F. McDermott, 2020). Our results show that only modest increases in treatment occur in response to E. coli. While informing households about contamination has been shown to increase treatment in some contexts (Jalan & Somanathan, 2008), the effectiveness of such informational interventions varies widely depending on design and context of the study (Davis, Pickering, Rogers, Mamuya, & Boehm, 2011; Okyere, Pangaribowo, & Gerber, 2019; Trent et al., 2018).

Second, we provide observational evidence on the relationship between household water treatment and drinking water quality. Although systematic reviews conclude that point-of-use water treatment is highly effective (Arnold, Colford Jr, et al., 2007; Fewtrell et al., 2005; Wolf et al., 2022), its real-world effectiveness is often reduced due to challenges in consistent use and unsanitary storage conditions (Clasen, Schmidt, Rabie, Roberts, & Cairncross, 2007; Sara V. Flanagan, Robert G. Marvinney, Robert A. Johnston, Qiang Yang, & Yan Zheng, 2015). Randomized controlled trials in Bangladesh, Kenya, and Zimbabwe show mixed impacts of water treatment on health outcomes, emphasizing the importance of adherence (Luby et al., 2018; Pickering et al., 2019). Our findings align with this literature, showing that while treatment reduces contamination, achieving fully safe drinking water requires factors more than simple access to water treatment methods.

Third, this study offers results with broad external validity, both across countries and within specific national and regional contexts. Water infrastructure and cultural practices around treatment vary widely by region and country. Using nationally representative samples from 25 countries, the study presents country-level statistics on water contamination, household water treatment, and the relationship between the two, helping to inform each country’s specific situation.

The remainder of this paper is organized as follows. Section 2 describes the data and methodology. Section 3 presents the empirical results. Section 4 concludes.

## 2 Methods

### 2.1 Sample

To examine the relationship between source water contamination, household water treatment decisions, and contamination in stored drinking water in the same framework, we analyze nationally representative household data from the MICS conducted in 25 countries across Africa, Latin America, and Asia.^2^ We accessed the data on September 3, 2024. Authors did not have access to any information that could identify individual participants during or after data collection. All MICS datasets are anonymized prior to public release. We use data collected between 2017 and 2021, in which a rapid water quality testing module is available. The surveys were implemented by UNICEF in collaboration with national ministries and statistics offices.

In households selected for water testing, interviewers requested permission to collect water samples from two locations: (1) the source from which the household collects water, and (2) the stored drinking water within the household. Interviewers first asked for a sample of drinking water by requesting a glass of drinking water, and then asked to be shown the water source location in order to collect a sample directly from the water source. These water samples were analyzed for the presence of *Escherichia coli* (E. Coli), an indicator of fecal contamination.^3^

Depending on the country, approximately 10–25 percent of a random subsample of households were selected for the rapid water quality testing module (Table A.1). Overall, 92.6 percent of households completed water testing with non-missing data, and in most countries, E. coli data are available for over 80–90 percent of the selected households. While non-consent was rare (N = 7), water testing was occasionally not conducted due to the household’s inability to provide a water sample, partial completion, or errors in reading the test results. A comparison of household characteristics between those who completed the water testing and those who did not shows varying patterns across countries (Appendix A). The final analytical sample includes 59,633 households from 25 countries.

### 2.2 Statistical Approach

This paper first presents the prevalence of four types of water treatment methods by country: (1) boiling, (2) use of chlorine products, Aquatabs, or PUR, (3) straining or settling, and (4) other methods. The water treatment methods analyzed refer specifically to the water treatment applied to the water sample for E. coli presence, which differs from the question on household water treatment practices in general.^4^

The study then presents the E.coli contamination levels of water at the source by country. E. coli levels are categorized based on the number of colony-forming units (CFUs) per 100 mL: less than 1, 1–100, and more than 100. For ease of description, this paper refers to water with less than 1 CFU as “low risk” or “free from E. coli,” 1–100 CFU as “moderate risk,” and more than 100 CFU as “high risk.”^5^

Water samples were collected from various types of sources, including piped water, tube wells and boreholes, protected wells or springs, unprotected wells or springs, surface or rainwater, and packaged or bottled water. Appendix C provides descriptive statistics, including contamination levels by source type. Contamination tends to be higher in water from wells, springs, and surface or rainwater sources.

Next, we conduct an empirical analysis to examine how household water treatment decisions vary with the E. coli contamination level at the water source. The analysis is performed using the pooled sample as well as disaggregated by region—Africa, Asia, and Latin America—to capture regional differences. We also assess whether specific treatment methods are associated with contamination levels.

The analysis employs a linear probability model to estimate binary outcomes of water treatment, allowing for interpretation of results in terms of percentage point changes in probability. All specifications include controls for country fixed effects, an urban residence indicator, water source type, and household socioeconomic status as defined by the MICS based on the asset index. Standard errors are clustered at the primary sampling unit (PSU) level.

The paper then examines contamination levels in stored drinking water. First, the paper focuses on households that do not treat their water to assess the natural increase in E. coli contamination that occurs during water storage over time. It then compares E. coli levels in source water and stored drinking water across different treatment methods to examine whether households that treat their water are able to reduce or eliminate contamination.

## 3 Results

### 3.1 Household Water Treatment and E. coli Contamination at Water Sources

Figure 1 shows the prevalence of water treatment practices by country. Overall, water treatment rates are low across most countries. With the exception of Guinea-Bissau, Mongolia, and Vietnam, fewer than 25 percent of households report treating their water. In Guinea-Bissau, treatment is primarily done through straining and settling, while in Mongolia and Vietnam, boiling is the most commonly reported method.

**Figure 1:**
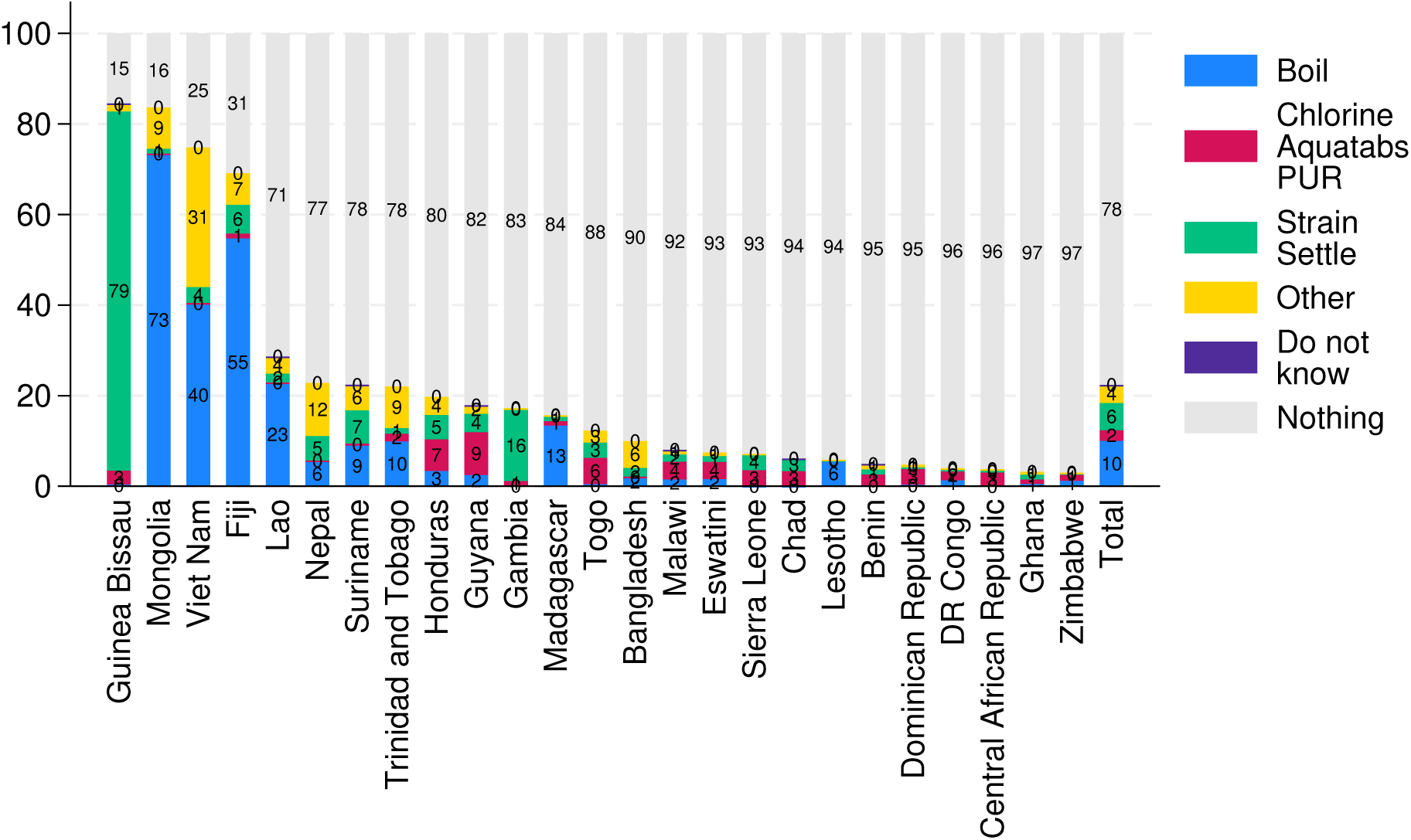
Water Treament Methods by Country. Notes: The sample is weighted, and the “Total” statistics are calculated as the average of country-level estimates. Other water treatment methods include options such as commercially available water filters.

Figure 2 (a) reports the E. coli contamination levels at the water source across countries. Countries such as Chad, Sierra Leone, and Madagascar exhibit high levels of contamination (*>*100 CFU/100ml). These countries not only have a greater share of households exposed to high-risk water sources, but also a substantial proportion with sources with moderate E. coli levels (1–100 CFU/100ml), resulting in less than 20 percent of households having access to low-risk water. Lesotho, Mongolia, and Trinidad and Tobago are characterized by relatively low levels of E. coli contamination, with over 70 percent accessing low-risk sources.

While overall water treatment rates is low, the critical measure is the treatment rate among households exposed to contaminated source water. Figure 2(b) builds on panel (a) by decomposing the share of households exposed to moderate or high risk into those that treat their water and those that do not.

**Figure 2:**
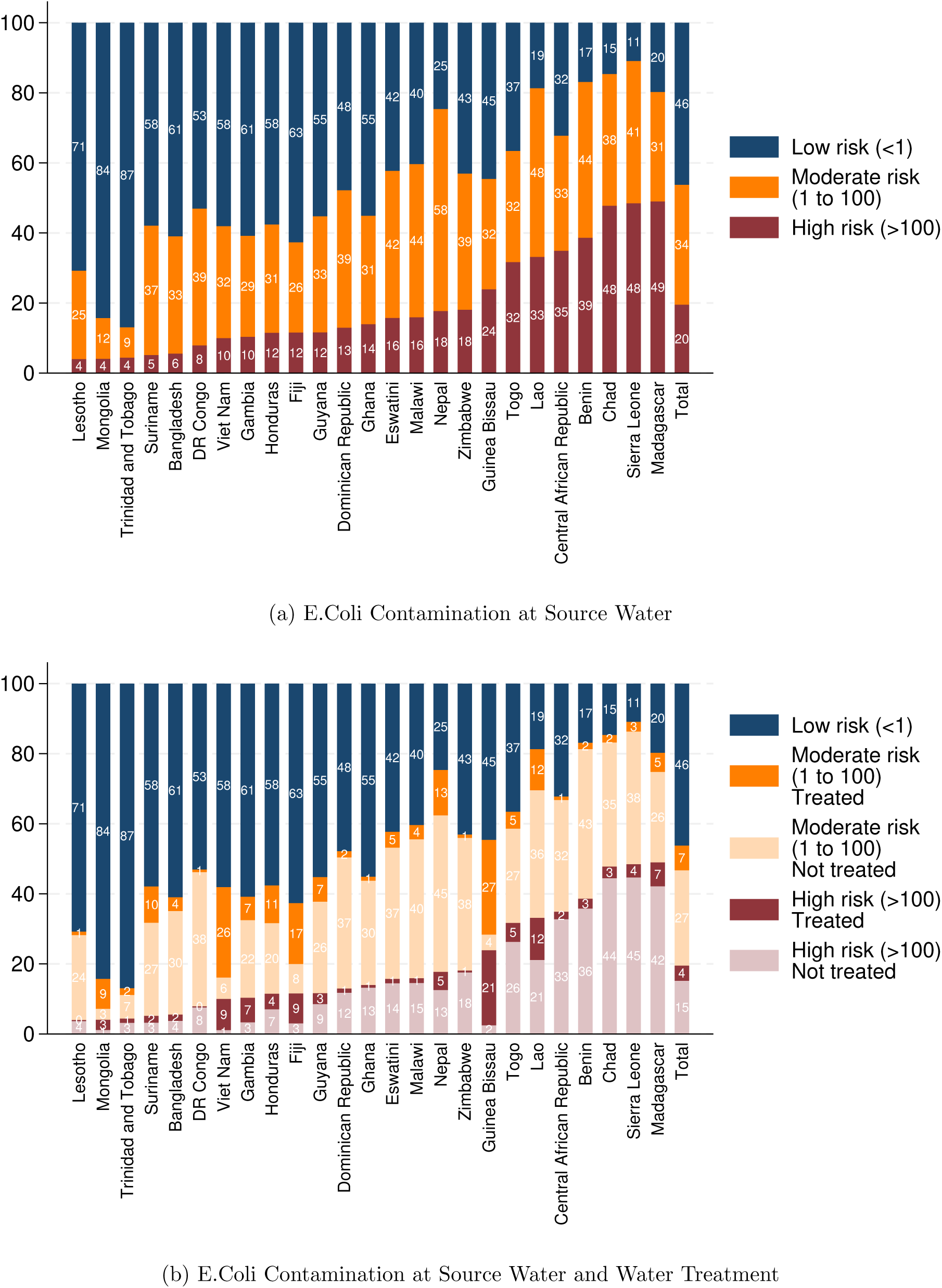
Water Source Contamination and Household Water Treatment by Country. Notes: Water treatment methods include boiling, chlorine products, Aquatabs or PUR, straining or settling, and other methods. The sample is weighted, and the “Total” statistics are computed as the average of country-level estimates.

Madagascar, Sierra Leone, and Chad shows a large proportion of households rely on water sources with high levels of E. coli contamination, while the prevalence of water treatment remains low. In Madagascar, 42 percent of households do not treat their water despite sourcing it from highly contaminated sources.

In Lesotho, although a relatively small share of households is exposed to moderate to high-risk water sources, overall water treatment rates are low, leading to 24 percent being exposed to moderate-risk water without any treatment.

In contrast, countries such as Mongolia, Vietnam, and Fiji show a low proportion of households exposed to highly contaminated water without treatment, reflecting their relatively high overall treatment rates. A similar pattern is observed in Guinea-Bissau, though this is primarily driven by the common use of straining and settling as a treatment method.

### 3.2 Water Treatment Decision in Relationship to Source Water Contamination

Table 1 presents the empirical results on how decisions to treat water differ by the contamination level of the water source. Column (1) shows that the probability of applying treatment is three percentage points higher when the water source has some presence of E. Coli compared to no presence. The probability increases to five percentage points if the water is highly contaminated.

**Table 1:** Water Source E.Coli Contamination on the Probability of Water Treatment.

|  | (1) | (2) | (3) | (4) | (5) | (6) | (7) | (8) |
| --- | --- | --- | --- | --- | --- | --- | --- | --- |
| Sample | Overall | Africa | Latin<br>America | Asia | Overall |  |  |  |
| Dependent var | Water treated (yes = 1, no = 0) |  |  |  | Boil | Chlorine<br>Aquatabs<br>PUR | Strain<br>Settle | Other |
| Base: Low risk |  |  |  |  |  |  |  |  |
| Moderate to high risk | 0.03***<br>(0.00) | 0.02***<br>(0.00) | 0.06***<br>(0.01) | 0.02**<br>(0.01) | -0.00<br>(0.00) | 0.01***<br>(0.00) | 0.02***<br>(0.00) | 0.00<br>(0.00) |
| Very high risk | 0.05***<br>(0.01) | 0.03***<br>(0.01) | 0.08***<br>(0.01) | 0.05***<br>(0.01) | -0.00<br>(0.00) | 0.02***<br>(0.00) | 0.03***<br>(0.00) | 0.01***<br>(0.00) |
| Country FE | Yes | Yes | Yes | Yes | Yes | Yes | Yes | Yes |
| Controls | Yes | Yes | Yes | Yes | Yes | Yes | Yes | Yes |
| Mean | 0.21 | 0.12 | 0.22 | 0.37 | 0.10 | 0.02 | 0.05 | 0.04 |
| Adjusted $R^2$ | 0.40 | 0.36 | 0.22 | 0.48 | 0.39 | 0.04 | 0.39 | 0.12 |
| Observations | 59,633 | 29,893 | 12,258 | 17,482 | 59,633 | 59,633 | 59,633 | 59,633 |

Note: All regressions control for country fixed effects, water source type, urban or rural location, and household socioeconomic status based on an asset-based wealth index. Standard errors are clustered at the primary sampling unit level (in parentheses). Significance levels are indicated as follows: \* $p < .10$ , \*\* $p < .05$ , \*\*\* $p < .01$ . \*

The presence of E. coli in source water is associated with a higher likelihood of household water treatment in all regions (Columns, 2–4). However, the increase is smaller in Sub-Saharan Africa. The largest effect is observed in Latin America, where E. coli contamination is associated with a six–eight percentage point increase in water treatment.

Columns (5)–(8) examine how the use of specific water treatment methods varies with the level of source water contamination. No significant increase in treatment is observed for boiling. However, chlorination methods—including Aquatabs and PUR—as well as straining and settling, show positive and statistically significant associations with higher contamination levels.

Table C.2 presents empirical results on water treatment by source type. However, because water treatment behavior and source selection involve complex decision-making processes that fall outside the scope of this paper, detailed estimations by source type are reported in the Appendix.

### 3.3 Household Water Treatment and E. coli Contamination in Drinking Water

Figure 3 (a) presents the levels of E. coli contamination in household stored drinking water. Compared to contamination at the source, drinking water shows higher levels of contamination. In Mongolia, 83 percent of households consume low risk drinking water. In contrast, in Chad, only one percent consume low-risk drinking water, while 77 percent consume water with high levels of contamination.

**Figure 3:**
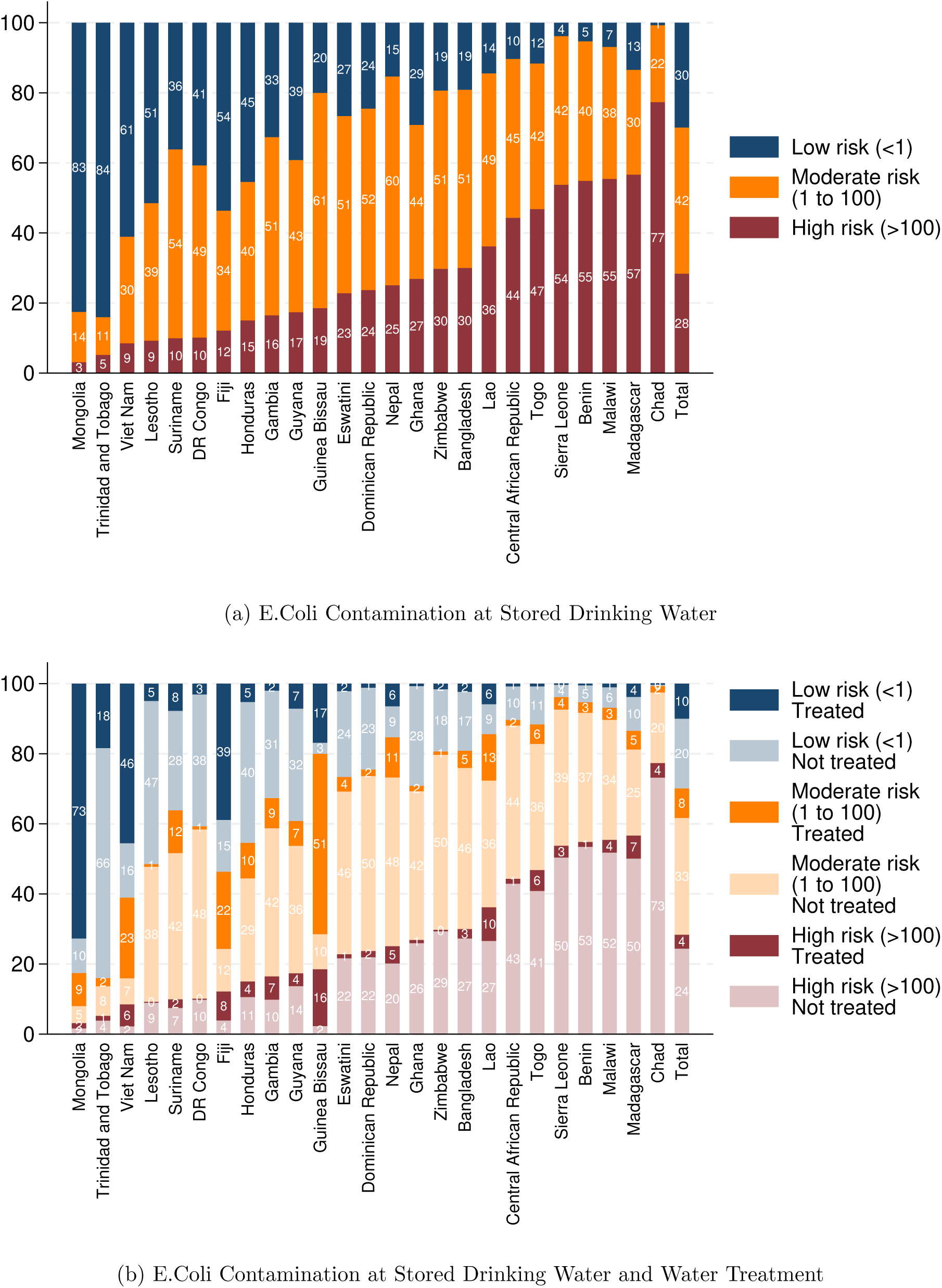
Stored Drinking Water Contamination and Household Water Treatment by Country. Notes: Water treatment methods include boiling, chlorine products, Aquatabs or PUR, straining or settling, and other methods. The sample is weighted, and the “Total” statistics are computed as the average of country-level estimates.

Figure 3 (b) reports a decomposition of drinking water contamination levels based on whether households treat their water. In Mongolia and Vietnam, where water treatment—particularly boiling—is widely practiced, a high share of households achieve low-risk drinking water. In contrast, in Guinea-Bissau, despite high reported treatment rates, which are primarily through straining and settling, 51 percent are still exposed to moderately contaminated drinking water. Additionally, 16 percent in Guinea-Bissau are exposed to high-risk drinking water despite treating it, suggesting limited effectiveness of the treatment methods used.

Figure 4 presents the transition of water contamination levels between the water source and stored drinking water. For households that do not treat their water, the share of households classified as low risk drops from 43 percent at the source to 22 percent in stored drinking water, while the proportions in moderate and high-risk categories increase.^6^

**Figure 4:**
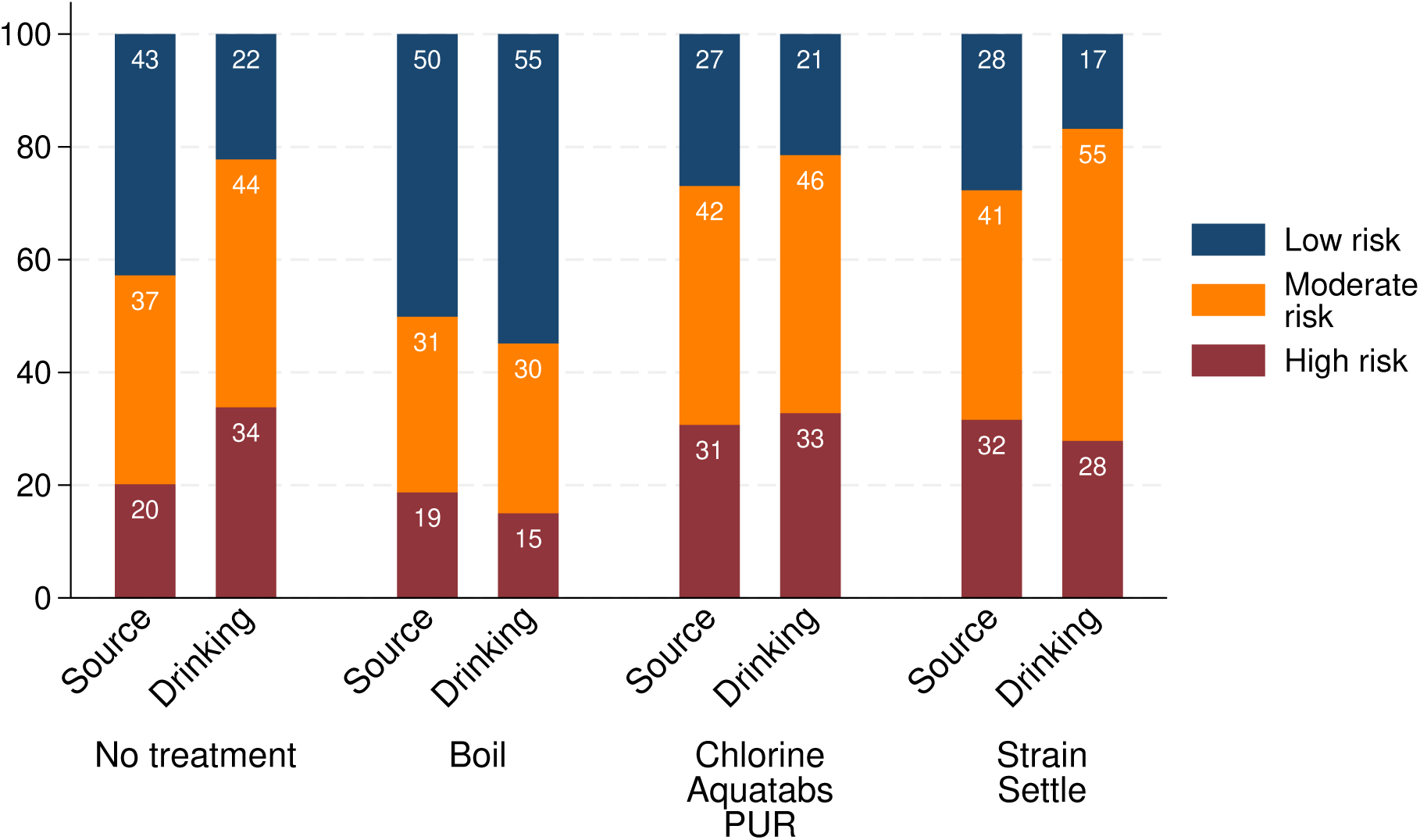
Water Contamination Levels in Source and Stored Drinking Water.

For any of the water treatment methods—boiling, chlorine-based products, or straining and settling—the overall distribution of contamination levels changes only modestly from source to drinking water. Among those who boil water, 19 percent of source samples are classified as high risk, decreasing slightly to 15 percent in stored drinking water. This suggests some reduction in contamination from boiling, but also underscores that boiling does not guarantee E. coli–free drinking water. Similar patterns are observed for chlorine-based treatments and straining/settling.

It is important to note that these results do not imply causal effects. Households’ choice of treatment method is closely linked to initial water quality and the surrounding environment, including storage conditions. Although water treatment methods such as boiling and chlorine-based products are effective at eliminating E. coli under controlled conditions, water can become recontaminated during storage in real-world settings. In addition, improper application of chlorine or the use of highly turbid or heavily contaminated water may require higher doses than households typically use. While the analysis in this paper is descriptive, the results from observational data highlight that a substantial share of households continue to consume water containing E. coli, even when they report using treatment methods.

## 4 Conclusion

Using data from 14 Sub-Saharan African countries, six Latin American countries, and five Asian countries, this paper documents E. coli contamination in source water and examines household responses to this invisible risk through water treatment practices. It also explores the relationship between these treatment practices and reductions in E. coli levels in stored drinking water.

Our analysis shows that only a small percentage of households treat their water, even when facing high contamination risks from E. coli in the water source, with levels exceeding 100 MPN/100ml. We find that as contamination levels increase, households are more likely to treat their water; however, the magnitude of this response is modest—ranging from three to five percentage points depending on the level of E. coli contamination. The response varies across regions and is primarily driven by increased use of chlorine, Aquatabs, PUR, and methods such as straining and settling, while no significant change is observed for boiling.

Even when households respond to contamination with water treatment, they often do not achieve E. coli-free water. These findings show the need for more robust and effective WASH interventions, specifically designed for regions with persistently low rates of water treatment adoption. Addressing these gaps is crucial for reducing the prevalence of waterborne diseases like diarrhea and saving lives across the region (Wolf et al., 2023).

Despite highlighting the link between water treatment and E. Coli levels, the paper has several limitations. One such limitation is that we do not make any causal claims. Instead, the paper focuses on the reduction of E. Coli by leveraging the availability of objective water quality measures, which are not typically included in household surveys. Another limitation is that the MICS data rely on self-reported information for water treatment practices, which is subject to social desirability bias. Respondents may over-report their adherence to water treatment methods, potentially leading to overestimating water treatment behavior. If the bias exists, the already concerning results of low water treatment practices may be even more worse thant what is reported in this paper.

## Data Availability

Data are available from the MICS Unicef website. It can be accessed from the following website: https://mics.unicef.org/surveys?display=card&f[0]=round:53

## Contributors

AK conceptualized the study, conducted the literature review, analyzed and interpreted the data, and drafted the original manuscript. BSS downloaded, cleaned, and pooled the data, contributed to the conceptualization of the study, conducted the literature review, and interpreted the data.

All authors reviewed and edited the manuscript. All authors had full access to all the data in the study. AK had final responsibility for the decision to submit for publication.

## Declaration of interests

All authors declare no competing interests.

## Data sharing

All data used for this and analytical codes are available immediately following publication without an end date to anyone for any purpose and are either published in the appendices or can be accessed through the corresponding author.

## Acknowledgments

We acknowledge the helpful comments from Rebecca Thornton, Jeremy Lowe, Elisa Maffioli, and Ana Radu.

## Appendices

### A Sample

Table A.1 reports the total number of households in the full survey, the number selected for the water quality testing module, and the number that provided consent. It also shows the number of samples with missing values for either water source or stored drinking water quality data, the number of samples missing water treatment information, and the final number of samples included in the analysis.

In most countries, E. coli water quality data are available for over 80-90 percent of households randomly selected for the water module. The total completion rate of the water testing with non-missing information is 92.6 percent. While non-consent for this module was low (N=7), water testing was often not conducted because households were unable to provide water or the testing was only partially completed. Honduras, Dominican Republic, and Suriname have the lowest completion rates, at 78.7 percent, 81.3 percent, and 81.5 percent, respectively.

Comparing household characteristics between those with and without water testing data across the full sample captures differences in completion rates at the country level, rather than differences within countries. For example, in Honduras, 1,088 households are missing water testing information. As a result, households with missing data in the full sample will disproportionately reflect the characteristics of Honduras, rather than revealing within-country differences.

Tables A.2–A.4 present household characteristics by testing completion status for Honduras, Suriname, and the Dominican Republic—the three countries with the lowest completion rates. Each country shows distinct patterns in the characteristics of households with missing water sample information. In Honduras, households with piped water are less likely to have complete data, while those using packaged or bottled water show higher completion rates. In contrast, this pattern is not observed in Suriname, where missing water sample data are more concentrated in urban areas and appear less related to the type of primary water source other than surface or rain water.

**Table A.1:**
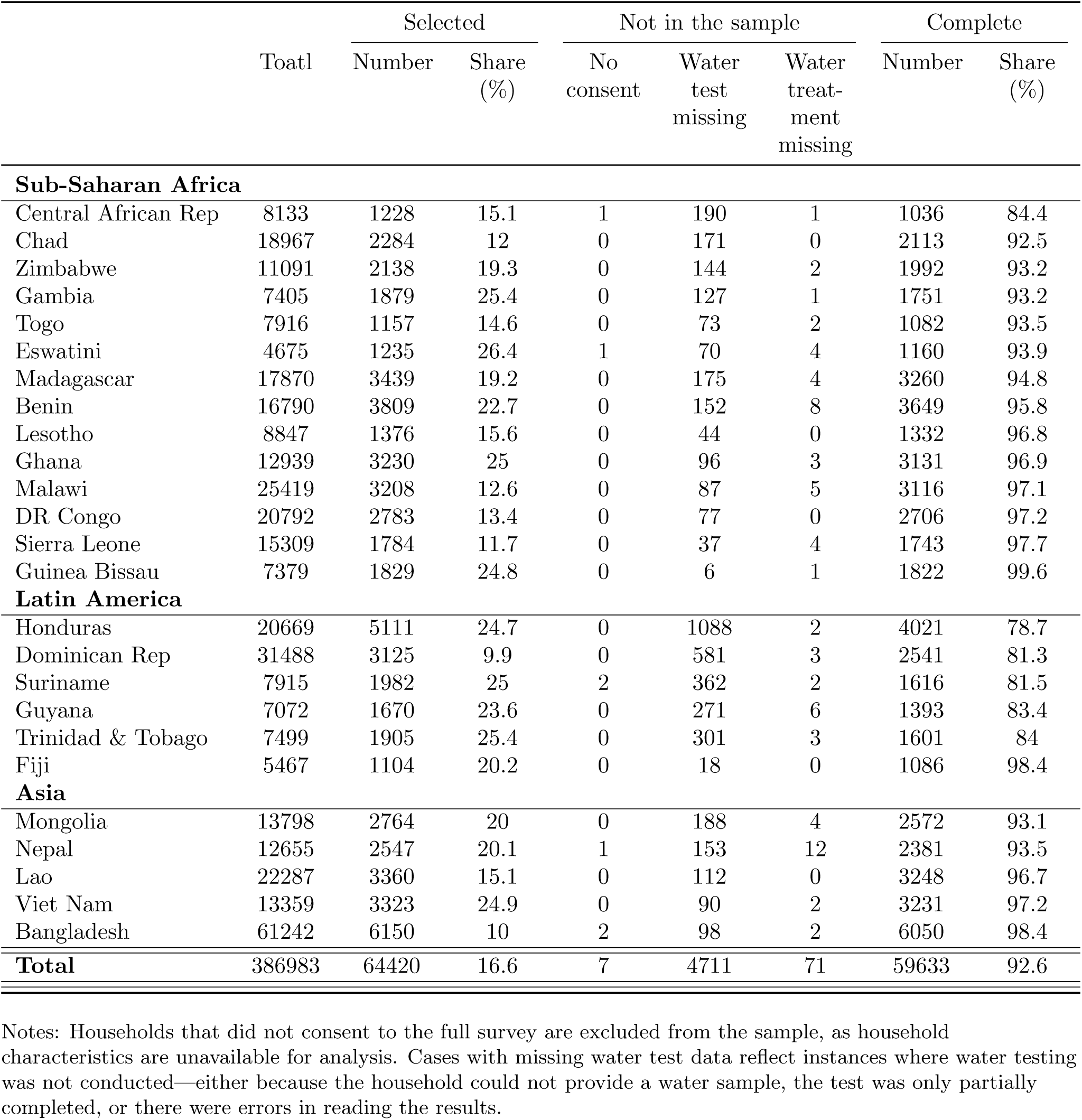
Number of Household Surveys and Sample Included in the Analysis.

**Table A.2:**
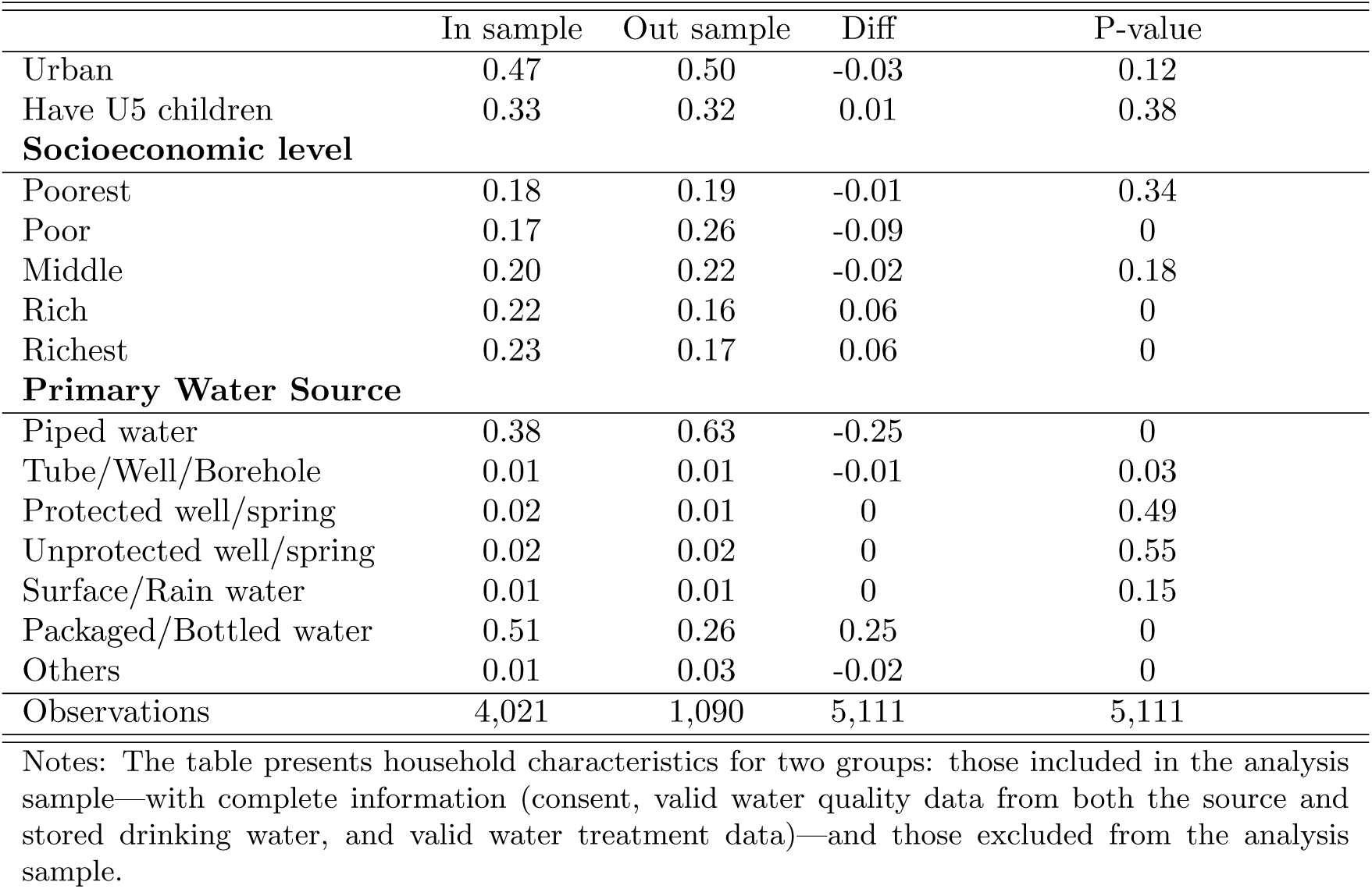
Household Characteristics for In/out Final Sample (Honduras)

**Table A.3:** Household Characteristics for In/out Final Sample (Suriname)

|  | In sample | Out sample | Diff | P-value |
| --- | --- | --- | --- | --- |
| Urban | 0.78 | 0.90 | -0.12 | 0 |
| Have U5 children | 0.25 | 0.23 | 0.02 | 0.33 |
| **Socioeconomic level** |  |  |  |  |
| Poorest | 0.20 | 0.18 | 0.02 | 0.43 |
| Poor | 0.18 | 0.22 | -0.03 | 0.14 |
| Middle | 0.22 | 0.21 | 0.01 | 0.54 |
| Rich | 0.21 | 0.14 | 0.07 | 0 |
| Richest | 0.19 | 0.25 | -0.06 | 0 |
| **Primary Water Source** |  |  |  |  |
| Piped water | 0.70 | 0.72 | -0.02 | 0.40 |
| Tube/Well/Borehole | 0 | 0 | 0 | 0.32 |
| Protected well/spring | 0.02 | 0.03 | -0.01 | 0.15 |
| Unprotected well/spring | 0 | 0 | 0 | 0.97 |
| Surface/Rain water | 0.17 | 0.11 | 0.06 | 0 |
| Packaged/Bottled water | 0.09 | 0.11 | -0.02 | 0.35 |
| Others | 0.01 | 0.02 | -0.02 | 0 |
| Observations | 1,616 | 366 | 1,982 | 1,982 |

Notes: The table presents household characteristics for two groups: those included in the analysis sample—with complete information (consent, valid water quality data from both the source and stored drinking water, and valid water treatment data)—and those excluded from the analysis sample.

**Table A.4:** Household Characteristics for In/out Final Sample (DominicanRepublic)

|  | In sample | Out sample | Diff | P-value |
| --- | --- | --- | --- | --- |
| Urban | 0.76 | 0.74 | 0.02 | 0.27 |
| Have U5 children | 0.24 | 0.19 | 0.05 | 0.01 |
| **Socioeconomic level** |  |  |  |  |
| Poorest | 0.22 | 0.26 | -0.05 | 0.01 |
| Poor | 0.20 | 0.23 | -0.03 | 0.07 |
| Middle | 0.20 | 0.19 | 0.01 | 0.43 |
| Rich | 0.20 | 0.20 | 0 | 0.94 |
| Richest | 0.19 | 0.12 | 0.06 | 0 |
| **Primary Water Source** |  |  |  |  |
| Piped water | 0.07 | 0.06 | 0.01 | 0.33 |
| Tube/Well/Borehole | 0.01 | 0 | 0 | 0.16 |
| Protected well/spring | 0 | 0.01 | -0.01 | 0.10 |
| Unprotected well/spring | 0 | 0 | 0 | 0.31 |
| Surface/Rain water | 0.02 | 0.03 | -0.01 | 0.23 |
| Packaged/Bottled water | 0.81 | 0.62 | 0.19 | 0 |
| Others | 0.01 | 0.01 | -0.01 | 0.20 |
| Observations | 2,541 | 584 | 3,125 | 3,125 |

Notes: The table presents household characteristics for two groups: those included in the analysis sample—with complete information (consent, valid water quality data from both the source and stored drinking water, and valid water treatment data)—and those excluded from the analysis sample.

### B Tables and Figure

**Figure B.1:**
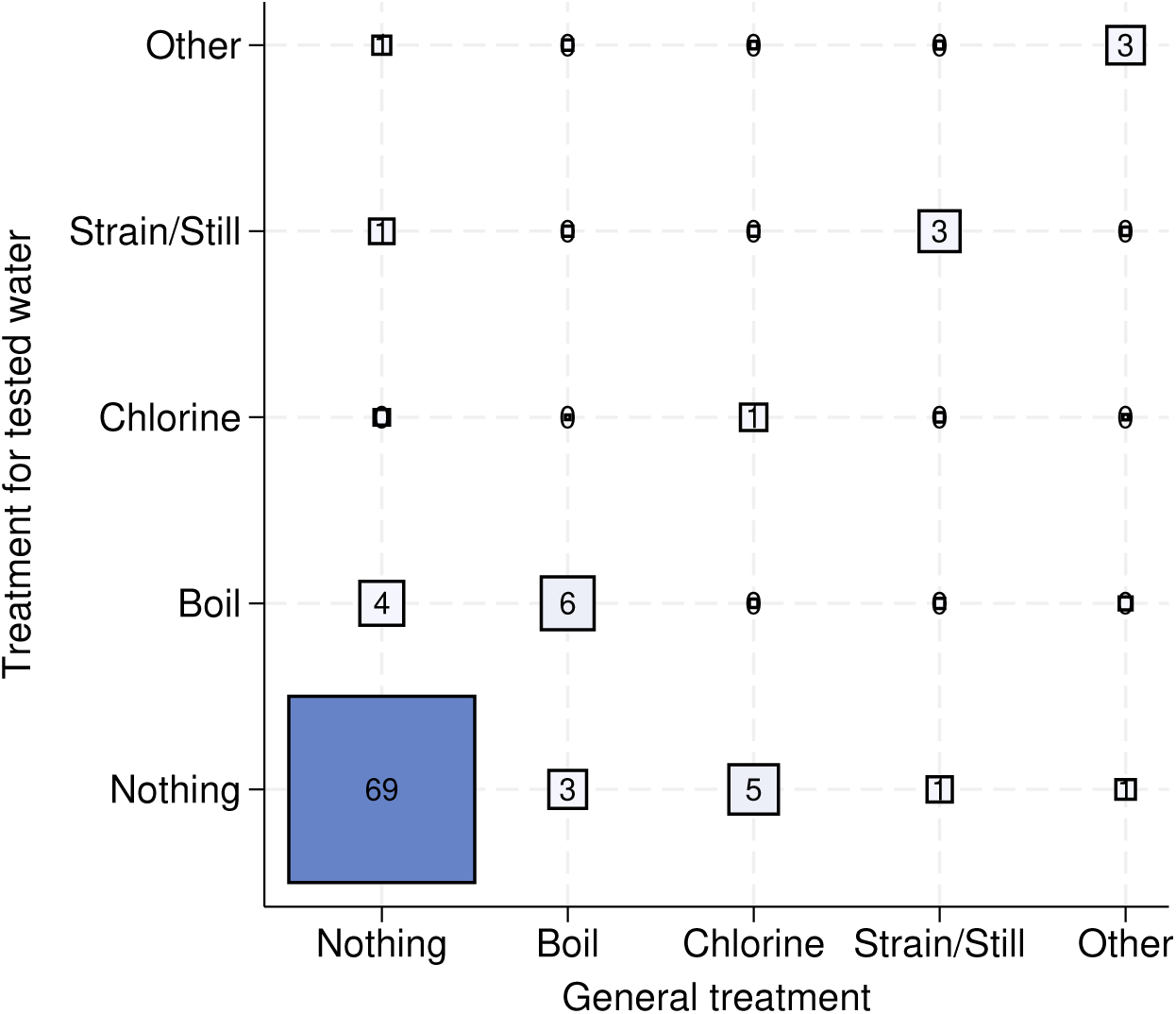
Water Treatment Methods for Water Tested for Quality and General Water Treatment

**Figure B.2:**
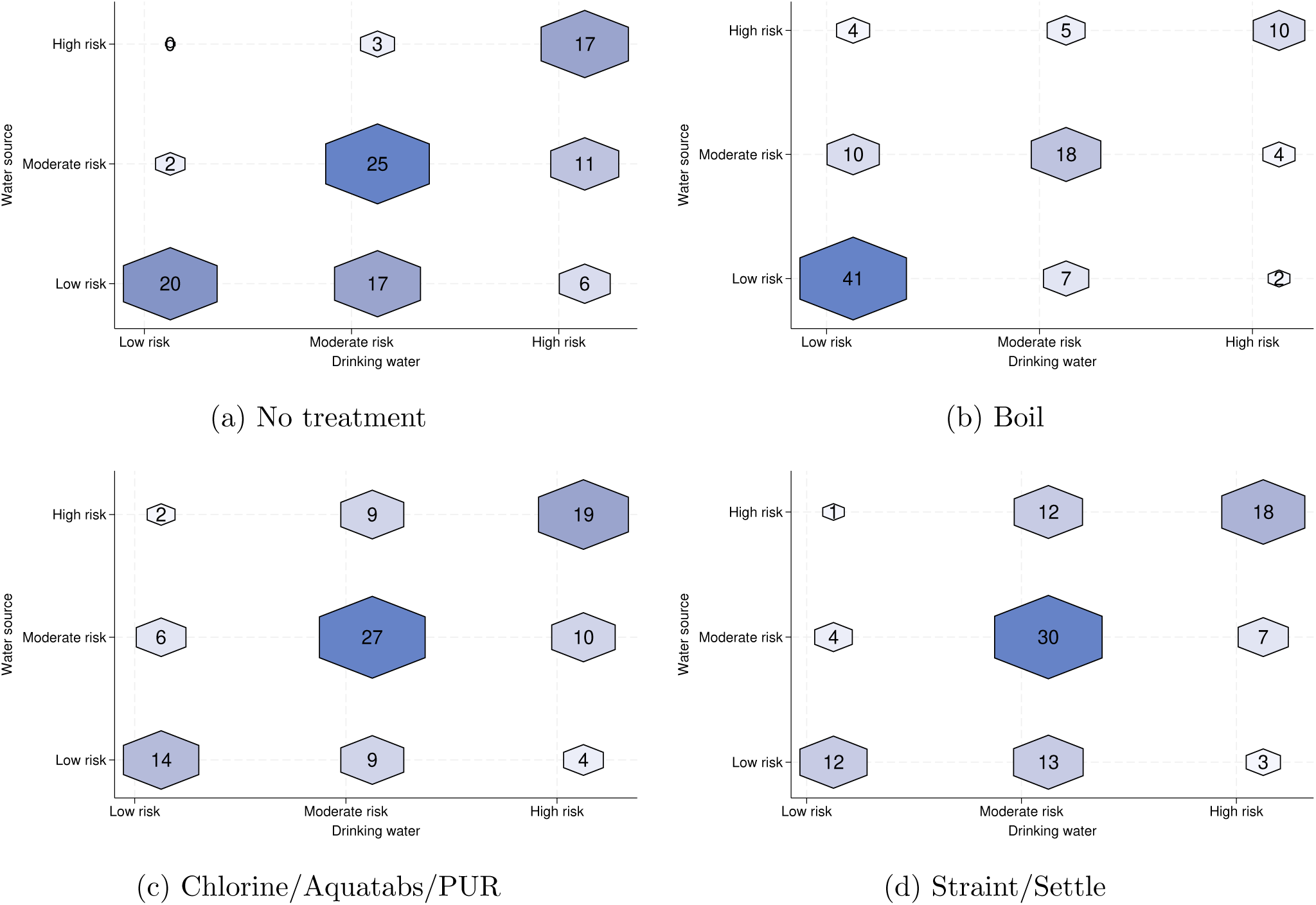
Transition of E. coli Contamination from Source to Drinking Water (percent)

### C Descriptive Statistics

Table C.1 presents household characteristics and water treatment behavior based on the contamination level of the source water. The final sample size is 59,633 households from 25 countries, with 25,518 (43 percent) in the low-risk category, 21,844 (37 percent) in the moderate-risk category, and 12,271 (21 percent) in the very high-risk category at the water source.

**Table C.1:** Household Characteristics by the Level of Water Source Contamination.

|  | Low risk | Medium risk | High risk |
| --- | --- | --- | --- |
| Urban | 0.55 | 0.35 | 0.24 |
| **Socioeconomic level** |  |  |  |
| Poorest | 0.13 | 0.23 | 0.31 |
| Poor | 0.16 | 0.21 | 0.24 |
| Middle | 0.20 | 0.20 | 0.20 |
| Rich | 0.23 | 0.20 | 0.15 |
| Richest | 0.28 | 0.17 | 0.10 |
| **Primary Water Source** |  |  |  |
| Piped water | 0.41 | 0.30 | 0.17 |
| Tube/Well/Borehole | 0.27 | 0.28 | 0.12 |
| Protected well/spring | 0.03 | 0.09 | 0.14 |
| Unprotected well/spring | 0.02 | 0.11 | 0.30 |
| Surface/Rain water | 0.02 | 0.07 | 0.18 |
| Packaged/Bottled water | 0.21 | 0.14 | 0.07 |
| Others | 0.03 | 0.01 | 0.01 |
| Any treatment (Water tested) | 0.23 | 0.20 | 0.22 |
| Nothing | 0.77 | 0.80 | 0.78 |
| Boil | 0.12 | 0.08 | 0.08 |
| Chlorine/Aquatabs/PUR | 0.01 | 0.03 | 0.03 |
| Strain/Settle | 0.03 | 0.06 | 0.08 |
| Other | 0.05 | 0.04 | 0.03 |
| **Water test results** |  |  |  |
| Source water test (100ml) | 0 | 21.10 | 101 |
| Household water test (100ml) | 17.80 | 44.26 | 86.12 |
| Observations | 25,518 | 21,844 | 12,271 |

Notes: The table presents the household characteristics across 25 countries. The number of CFUs per 100mL of E. Coli is capped at 101 if the number is higher than 100. The mean of the blank water test is 0.67 CFUs per 100mL.

Source water contamination is linked to urbanization: urban households make up 55 percent of the no-risk category, 35 percent of the moderate-risk category, and 24 percent of the high-risk category. Additionally, higher socioeconomic status is strongly associated with lower contamination levels in drinking water.

Water contamination is closely linked to the source, with 54 percent of uncontaminated water coming from piped sources, while only 11 percent of very high-risk water originates from piped sources. Very high-risk water is predominantly found in unprotected wells, springs, and surface water sources, including rainwater. Water treatment rates are low overall, and when households do treat their water, boiling, chlorination, straining, or letting water settle are the most common methods.

The empirical analysis shows higher treatment rates in response to higher levels of source contamination only for households using piped water, as well as packaged and bottled water.

**Table C.2:** Water Source E.Coli Contamination on the Probability of Water Treatment.

|  | Dependent var: Water treated (yes = 1, no = 0) |  |  |  |  |  |  |
| --- | --- | --- | --- | --- | --- | --- | --- |
|  | (1) | (2) | (3) | (4) | (5) | (6) | (7) |
|  | Overall | Piped water | Tube Well Borehole | Protected well spring | Unprotected well spring | Surface Rainfall | Packaged Bottle |
| Base: Low risk |  |  |  |  |  |  |  |
| Moderate to high risk | 0.03***<br>(0.00) | 0.03***<br>(0.01) | 0.00<br>(0.00) | -0.00<br>(0.02) | 0.00<br>(0.01) | -0.02<br>(0.02) | 0.03***<br>(0.01) |
| Very high risk | 0.05***<br>(0.01) | 0.08***<br>(0.01) | 0.01<br>(0.01) | 0.00<br>(0.02) | -0.02<br>(0.01) | -0.02<br>(0.02) | 0.02**<br>(0.01) |
| Country FE | Yes | Yes | Yes | Yes | Yes | Yes | Yes |
| Controls | Yes | Yes | Yes | Yes | Yes | Yes | Yes |
| Mean | 0.21 | 0.30 | 0.25 | 0.08 | 0.13 | 0.08 | 0.14 |
| Adjusted $R^2$ | 0.40 | 0.39 | 0.47 | 0.51 | 0.53 | 0.27 | 0.09 |
| Observations | 59,633 | 17,987 | 14,836 | 4,823 | 7,501 | 4,769 | 8,566 |

Note: All regressions control for country fixed effects, water source type, urban or rural location, and household socioeconomic status based on an asset-based wealth index. Standard errors are clustered at the primary sampling unit level (in parentheses). Significance levels are indicated as follows: \* $p < .10$ , \*\* $p < .05$ , \*\*\* $p < .01$ . \*

## Footnotes

1 The overall estimates in this paper are calculated as simple averages of nationally representative statistics at the country level.

2 From an initial sample of 29 countries, we excluded Tuvalu, Kiribati, Tonga, and the Turks and Caicos Islands, where fewer than 1,000 households completed the water testing module, to ensure sufficient data for reliable country-level statistics.

3 The drinking water quality module for the survey was developed collaboratively by the Joint Monitoring Program (JMP) and the global MICS program. An international water quality trainer, with support from national experts, led the training of field teams. National experts from regulatory agencies, research labs, or ministries of health provided technical input and oversight during fieldwork through a few field visits. Water samples were analyzed for E. Coli using portable membrane filtration. Field teams filtered 100mL of water through a 0.45-*µ*m filter, placed it on CompactDry EC growth media plates, and rehydrated it with 1mL of sample water.

4 Figure B.1 shows the relationship between water treatment reported for the tested water sample and general water treatment practices. Treatment rates are lower for the tested water, suggesting that households may not consistently apply the treatment methods they report using in general. While households are more likely to report using chlorine in response to the general question, boiling is more frequently reported for the water sample that was tested.

5 World Health Organization (2017) use a more detailed classification, defining 1–10 CFU as moderate risk, 10–100 CFU as high risk, and more than 100 CFU as very high risk.

6 Figure B.2 presents a 3×3 transition matrix showing how contamination levels change from source to stored water. Among households without water treatment, only a small fraction improve their contamination category, indicating water contamination level stays in the same category or deteriorates without any water treatment.

## Notes

### Competing Interest Statement

The authors have declared no competing interest.

### Funding Statement

The author(s) received no specific funding for this work.

